# Association of genetic variants in the autophagy gene ATG4B with asthma

**DOI:** 10.64898/2026.03.19.26348866

**Authors:** Dyema Nicholson, Andrew T. DeWan

**Author notes:** To whom correspondence should be addressed: Dr. Andrew T. DeWan, Department of Chronic Disease Epidemiology, Center for Perinatal, Pediatric and Environmental Epidemiology, Yale School of Public Health, 1 Church Street, 6^th^ Floor, New Haven, Connecticut, 06510, USA. **Funding** This work was funded by a grant from the National Institute of Environmental Health Sciences (R25ES029052). **Data Availability** Summary statistics were access through the GWAS Catalog (GCST90302886).

## Abstract

Asthma is a chronic respiratory illness that causes mild to severe inflammation throughout narrowed airways. During allergic airways inflammation, autophagy prevents extensive lung tissue impairment while inducing a protective anti-pathogen response and macrophages in the lung to maintain homeostasis. Previous studies of autophagy genes and asthma have shown an association with variants in ATG5, but a comprehensive analysis of autophagy related genes and asthma has not been performed. Here we utilize summary statistic data generated from a two-stage genome-wide association study (GWAS) of asthma in the UK Biobank. We examined variants in 21 autophagy related genes and found statistically significant associations for 28 variants in two genes in the discovery dataset and nominally significant replication for 16 of these variants, all annotated to ATG4B. This is the first evidence of an association with variants in ATG4B with asthma which provides a novel potential for future drug development.

## Introduction

Asthma, affecting approximately 262 million people worldwide,(1) is a chronic respiratory illness that causes mild to severe inflammation throughout narrowed airways. There are many factors that contribute to the development of asthma including pregnancy-related factors (e.g. smoking during pregnancy, mode of delivery),(2,3,4,5,6) environmental factors (second hand smoke exposure, indoor and outdoor air pollution),(7,8,9) and genetic factors.(10) Although asthma is multi-factorial in nature, it is believed that one common cause of asthma is contact of an allergen with an altered immune system leading to inflammation.(11) During allergic airway inflammation, autophagy prevents extensive lung tissue impairment while inducing a protective anti-pathogen response and macrophages in the lung to maintain homeostasis.(12)

Autophagy includes the process of cytoplasmic self-degradation and the recycling of cytoplasmic organelles in lysosomes to regulate and maintain homeostasis and promote immunity.(13) It consists of an adaptive pathway that when induced by reactive oxygen species (ROS), has an active role in protecting cells by preventing the collection of damaged mitochondria and mitigating oxidative stress.(14) Pulmonary circulation is reliant on gas exchange, and so an over production of ROS or a mutation of an autophagy-related gene can lead to the development of asthma or related pulmonary disease.(15)

A previous study focused on genetic variants in two autophagy genes, ATG5 and ATG7, and found an association between variants in ATG5 and childhood asthma.(16) However, a comprehensive analysis of genetic variants a broader list of autophagy related genes has not be performed.

This paper aims to assess whether genetic variants annotated to genes in the autophagy pathway are associated with the development of asthma.

## Methods

### Data access and ethical approval

This research was conducted using the UK Biobank Resource (application number 32285). The UK Biobank study was conducted under generic approval from the National Health Services’ National Research Ethics Service. The present analyses were approved by the Human Investigations Committee at Yale University (protocol number 2000026836).

The results presented here utilized the summary statistics generated from our previous paper (17) and are described below.

### UK Biobank Discovery Dataset

Participants were enrolled in the UK Biobank, and data accessed under an approved agreement (application ID 32285). Genotypes were assayed on either the UK BiLEVE array or the UK Biobank Axiom array with 733,332 autosomal variants overlapping between the two arrays. Subject and variant quality control is described in Supplemental Table S12, with subjects limited to those genetically defined as White British (field 22006) and with a call rate >99% and variants limited to those with a call rate >99%, HWE p-value > 5x10^-8^ and minor allele frequency (MAF) > 0.01. This resulted in 366,752 individuals and 529,024 directly genotyped variants for analysis. In addition, imputed variants obtained directly from the UK Biobank were included. Variants were filtered based on MAF > 0.001 and Info Score > 0.8 yielding 13,407,279 imputed variants.

### Replication Dataset

For a replication, we identified a set of White non-British individuals using self-reported ancestry and pre-computed principal components (PCs; field 22009). First, the mean and covariance was calculated for the 40 PCs for all subjects (N= 488,263). For each subject, the Mahalanobis Distance of the empirical PC distribution was calculated. Among all 488,263 UKB subjects, 409,615 are genetically confirmed white British subjects (field 21000); 50,518 are self-reported white European but not genetically confirmed white British subjects; and 28,130 are self-reported non-white subjects. Using a Mahalanobis Distance threshold that captures 94% (458,967) of subjects, we include all genetically confirmed white British subjects, 46,352 self-reported white European non-British subjects and 2,763 self-reported non-white subjects. The 46,352 subjects were then considered for the replication dataset. The same quality control parameters as for the discovery dataset were used (Supplemental Table S2). The final replication dataset consisted of 44,173 individuals and 13,793,916 imputed variants, of which 550,028 were directly genotyped.

### Phenotype Definitions

Asthma was defined by either ICD-10 code (field 41270, J45 or J46) or self-reported diagnosis by a doctor (field 6152). Individuals with autoimmune conditions were excluded from the controls (field 20002, self-reported diagnosis of an autoimmune disease (18) or self-report of sarcoidosis diagnosis by doctor in field 22133). In the discovery dataset, there were 48,623 cases and 290,722 controls. In the replication dataset, there were 5,822 cases and 36,615 controls.

### Principal Components Analyses

Using the directly genotyped variants, the data was linkage disequilibrium (LD) pruned using the default parameters of pruning variants in a 50 variant window so that none have a pairwise r2 of greater than 0.2 and shifting by 5 variants after each step in Plink-1.90 (19). Principal components analysis was performed for each trait separately using the smartPCA program in EIGENSOFT-7.2.1.(20)

### Univariate Analysis

Univariate association analysis was performed using linear and logistic regression models implemented in REGENIE.(21) REGENIE implements a multi-stage procedure which starts by using a whole genome regression strategy, closely related to the linear mixed model, using Ridge regression to estimate the polygenic effects parameter to account for relatedness and population stratification. For binary traits, Firth regression is used to ensure that the results are well calibrated in the presence of low frequency variants and unbalanced case-control ratios, both of which are present in this analysis. REGENIE implements an approximate Firth regression to ensure that this approach can be run on a genome-wide scale. The analyses were adjusted for age at recruitment (field 21022), genetic sex (field 22001), and 10 principal components.

### Autophagy genes

A set of 21 candidate genes in the autophagy pathway were selected due to their roles during autophagy and previous studies relating them to the development of asthma (Table 1).(22)(23) There were 17 autophagy genes selected (ATG2A through ATG16L1 in Table 1), along with microtubule-associated protein 1A/1B-light chain 3 (MAP1LC3A and MAP1LC3B) as they aid in the formation and elongation of autophagosomes and the two genes that comprise the ULK complex (ULK1 and ULK2) due to their influence on the cellular and molecular mechanisms during lysosome formation which is an early stage of autophagy.(24)(25)(26)

**Table 1.** Autophagy-related candidate genes.

| **Genes** | **Chr<sup>1</sup>** | **Start position (bp)<sup>2</sup>** | **Stop position (bp)<sup>2</sup>** | **Number of SNPs located (Discovery data)** |
| --- | --- | --- | --- | --- |
| ATG4C | 1 | 63249777 | 63330941 | 436 |
| ATG9A | 2 | 220082847 | 220094361 | 172 |
| ATG16L1 | 2 | 234160217 | 234204320 | 405 |
| ATG4B | 2 | 242577027 | 242613271 | 498 |
| ATG7 | 3 | 11314010 | 11599139 | 1372 |
| ATG3 | 3 | 112251359 | 112280485 | 365 |
| ATG10 | 5 | 81267844 | 81551216 | 1292 |
| ATG12 | 5 | 115163894 | 115177548 | 230 |
| ATG5 | 6 | 106632352 | 106773695 | 818 |
| ATG9B | 7 | 150709297 | 150721586 | 259 |
| ATG13 | 11 | 46638826 | 46697568 | 255 |
| ATG2A | 11 | 64662004 | 64684722 | 207 |
| ATG16L2 | 11 | 72525451 | 72540680 | 205 |
| ULK1 | 12 | 132379279 | 132407707 | 334 |
| ATG14 | 14 | 55833109 | 55878576 | 442 |
| ATG2B | 14 | 96747595 | 96829678 | 461 |
| MAP1LC3B | 16 | 87425801 | 87438380 | 299 |
| ULK2 | 17 | 19674143 | 19771239 | 416 |
| ATG6 | 17 | 40962766 | 40968011 | 68 |
| ATG4D | 19 | 10654647 | 10664094 | 309 |
| MAP1LC3A | 20 | 33134688 | 33148149 | 136 |

<sup>1</sup>Chr = Chromosome
<sup>2</sup>base pair position using hg19 coordinates

The transcription start and stop sites of these 21 genes were obtained from the UCSC Genome Browser (using GRCh37/hg19 coordinates). An additional 20KB up and downstream of each start and stop site, respectively, was added to each gene coordinate to capture potential cis-regulatory variants around each gene. These coordinates were then used to extract the association results for the SNPs annotated to each of the autophagy genes.

There were 8,979 SNPs mapped to the 21 genes. A p-value of 5.6 x 10-6 was used as the significant threshold to account for multiple testing (p=0.05/8,979). Significant variants from the discovery sample were examined in the replication sample. Replication was declared if the variants had a p<0.05 and a beta in the same direction in the replication sample.

## Results

A total of 28 variants were significant at p<5.6x10-6 in the discovery dataset (Table 2). Of these, 26 were in or around ATG4B. The remaining two variants were in or around ATG4D. The most significant variant, rs34143604, is 15,017 bp upstream of the ATG4B gene, within an intron of the gene THAP4. Of the 28 significantly associated variants, 16 were significantly associated at p<0.05 and had betas in the same direction in the replication dataset. All 16 variants were in or around ATG4B, but did not include the most significantly associated variant, rs34143604.

**Table 2.** Significant variants in autophagy candidate genes in discovery sample (p<5.6x10-6) and evidence for replication.

| Gene | Chr<sup>1</sup> | Variant<sup>2</sup> | Position<sup>3</sup> | ALLELE0 | ALLELE1 | A1FREQ | Discovery |  |  |  |  |
| --- | --- | --- | --- | --- | --- | --- | --- | --- | --- | --- | --- |
|  |  |  |  |  |  |  | INFO<sup>4</sup> | BETA | SE<sup>5</sup> | P<sup>6</sup> | INFO<sup>4</sup> |
| ATG4B | 2 | rs34143604 | 242562010 | A | C | 0.620 | 0.990 | -0.045 | 0.007 | 5.63E-10 | 0.988 |
| ATG4B | 2 | rs7599737 | 242569897 | A | T | 0.560 | 0.990 | -0.041 | 0.007 | 1.07E-08 | 0.989 |
| ATG4B | 2 | rs62191287* | 242625413 | T | A | 0.576 | 0.997 | 0.033 | 0.007 | 4.34E-06 | 0.996 |
| ATG4B | 2 | rs7369213* | 242625734 | G | C | 0.576 | 0.997 | 0.033 | 0.007 | 3.19E-06 | 0.996 |
| ATG4B | 2 | rs7370850 | 242625935 | C | G | 0.414 | 0.996 | 0.037 | 0.007 | 2.13E-07 | 0.995 |
| ATG4B | 2 | rs35925884* | 242626479 | T | C | 0.576 | 0.998 | 0.033 | 0.007 | 3.60E-06 | 0.998 |
| ATG4B | 2 | rs7597410 | 242626822 | A | G | 0.414 | 0.996 | 0.037 | 0.007 | 2.05E-07 | 0.995 |
| ATG4B | 2 | rs150059971* | 242626941 | TTTG | T | 0.589 | 0.971 | 0.035 | 0.007 | 1.62E-06 | 0.970 |
| ATG4B | 2 | 2:242626965* | 242626965 | C | CAG | 0.575 | 0.998 | 0.033 | 0.007 | 4.02E-06 | 0.997 |
| ATG4B | 2 | rs7371228* | 242627097 | C | G | 0.567 | 0.988 | 0.033 | 0.007 | 4.23E-06 | 0.987 |
| ATG4B | 2 | rs35655036* | 242627715 | TTC | T | 0.581 | 0.987 | 0.034 | 0.007 | 2.83E-06 | 0.987 |
| ATG4B | 2 | rs35416933* | 242627938 | C | T | 0.576 | 1.000 | 0.033 | 0.007 | 2.66E-06 | 1.000 |
| ATG4B | 2 | rs34424237* | 242628078 | G | C | 0.576 | 1.000 | 0.033 | 0.007 | 3.64E-06 | 1.000 |
| ATG4B | 2 | rs34836619* | 242628126 | T | C | 0.576 | 1.000 | 0.033 | 0.007 | 3.83E-06 | 1.000 |
| ATG4B | 2 | rs34302342* | 242628463 | G | A | 0.576 | 1.000 | 0.033 | 0.007 | 3.77E-06 | 0.999 |
| ATG4B | 2 | 2:242628534* | 242628534 | A | AT | 0.575 | 0.999 | 0.033 | 0.007 | 3.42E-06 | 0.999 |
| ATG4B | 2 | rs6437279* | 242628768 | A | G | 0.576 | 0.999 | 0.033 | 0.007 | 3.64E-06 | 0.999 |
| ATG4B | 2 | rs34343771 | 242629529 | A | G | 0.415 | 0.997 | 0.037 | 0.007 | 1.78E-07 | 0.996 |
| ATG4B | 2 | rs7573214 | 242630048 | A | G | 0.415 | 0.997 | 0.037 | 0.007 | 1.82E-07 | 0.996 |
| ATG4B | 2 | rs62191297 | 242630680 | C | A | 0.457 | 0.998 | 0.039 | 0.007 | 3.05E-08 | 0.997 |
| ATG4B | 2 | rs75447999 | 242630864 | C | A | 0.457 | 0.998 | 0.039 | 0.007 | 2.25E-08 | 0.997 |
| ATG4B | 2 | rs7574248* | 242630943 | A | G | 0.577 | 0.996 | 0.034 | 0.007 | 1.91E-06 | 0.995 |
| ATG4B | 2 | rs143872071 | 242630957 | CTT | C | 0.457 | 0.996 | 0.039 | 0.007 | 2.42E-08 | 0.994 |
| ATG4B | 2 | rs62191299* | 242630965 | A | G | 0.576 | 0.997 | 0.034 | 0.007 | 1.99E-06 | 0.996 |
| ATG4B | 2 | rs7574259* | 242630976 | C | G | 0.575 | 0.998 | 0.034 | 0.007 | 2.22E-06 | 0.997 |
| ATG4B | 2 | rs34174712 | 242631579 | C | T | 0.456 | 0.997 | 0.039 | 0.007 | 2.59E-08 | 0.996 |
| ATG4D | 19 | rs549956721 | 10636964 | G | A | 0.971 | 0.900 | -0.106 | 0.021 | 8.77E-07 | 0.859 |
| ATG4D | 19 | rs183143244 | 10665021 | C | A | 0.975 | 0.973 | -0.107 | 0.022 | 2.05E-06 | 0.967 |

<sup>1</sup>Chr = Chromosome
<sup>2</sup> Variants with evidence of replication ( $p < 0.05$ and Beta in the same direction) are indicated with \*
<sup>3</sup> base pair position using hg19 coordinates
<sup>4</sup>INFO = information score which indicates the quality of the imputation
<sup>5</sup>SE = standard error
<sup>6</sup>p = p-value

## Discussion

The ATG4 protein is necessary for cleaving C-terminal segments of proteins, performing lipid deconjugation which is required for target recycling, and developing autophagy-specific inhibitors, which all play a role in regulating autophagy.(27) ATG4 consists of four homologues including, ATG4A, ATG4B, ATG4C, and ATG4D. Based on our results, ATG4B and ATG4D were the only genes in the autophagy pathway that had variants that were significantly associated with asthma. While ATG4D contributes less to the catabolism of proteins and peptides, ATG4B contributes the most protease activity towards ATG8 homologues in generating MAP1LC3A and recycling MAP1LC3B.(28) The effect of ATG4 homologues on autophagy based upon their enzyme kinetics with ATG8 homologues explored in mammalian cell-based studies concluded that the manipulation of ATG4C and ATG4D produced minimal activities, while the ATG4B and ATG4A enzymes induced more notable results.(27)

Though there are limited studies that examine the relationship between ATG4 homologues with asthma, there are extensive studies investigating ATG4 homologues with the development of pulmonary diseases. One experimental study examined the effect of autophagy impairment in lung fibrosis *in vivo*.(29) Using ATG4B-deficient mice, this study was able to show that ATG4B was involved in the regulation of lung homeostasis, protection of epithelial cells against apoptosis, and has a role in inflammatory and fibrotic response during bleomycin-induced stress. However, the effect of autophagy deficiency on the development of lung fibrosis has yet to be observed.

Finding and implementing personalized treatment plans for asthma is the key to developing the next class of drugs that will reduce symptoms and ensure a good quality of life. Based on our findings of an association with genetic variants in and around ATG4B, ATG4-specific inhibitors(30) may be good candidates for drug development for asthma subtypes.

## Data Availability

Summary statistics are available in the GWAS Catalog (GCST90302886)

## Acknowledgements

We thank the Yale Center for Research Computing for the use of the McCleary High Performance Computing cluster.

